# Patterns of treatment and outcome in patients with unresectable or inoperable esophageal cancer: A real world data

**DOI:** 10.1101/2022.01.26.22269828

**Authors:** Richa Chauhan, Vinita Trivedi, Rita Rani, Usha Singh

## Abstract

**Background:** Esophageal cancer is the eighth most common cancer in the world with a high mortality rate. Surgery, radiation and chemotherapy have been tried in various combinations to improve on the survival rates. Our study provides real world data from a South Asian country on patterns of treatment and outcome in patients with unresectable or inoperable esophageal cancer.

**Materials and Methods:** This study is a retrospective analysis of all consecutive esophageal cancer patients, with medically inoperable or unresectable disease, and treated with conformal radical radiotherapy at a tertiary cancer center from January, 2016 to December, 2017. Data regarding patients’ age, histology, location, pre-treatment imaging, disease stage, treatment details, compliance and response to treatment and status at last follow-up were retrieved from their file. Continuous and categorical variables were summarized by descriptive statistics.

**Results:** A total of 100 esophageal cancer patients with a mean age of 60.24 years were included in the study. 60% of the patients were male and upper one-third was the most common site involved. Squamous cell carcinoma was reported in 83% of the patients. About 70% of the patients had a T3/T4 disease and 44% also had nodal metastasis. The radiation dose ranged from 45Gy – 63Gy (median = 59.4Gy). Further, 15% and 54% of the patients received neoadjuvant and concurrent chemotherapy respectively. Radiation compliance was seen in 90% of the patients. With a median follow-up of 7 months (range 3-58 months), 80% of the patients were alive with 32.22% having no evidence of disease. Distant metastases and loco regional failure was seen in 32.22% and 28% of the patients respectively.

**Conclusion:** Our study showed that esophageal cancer is more common in elderly males. Adherence to a uniform treatment protocol using concurrent chemo radiation is difficult in clinical practice especially in resource constrained set up. Both distant metastases and loco regional failure continues to be a matter of concern. Further improvement in local control must be evaluated by either radiation dose escalation or novel combinations with chemotherapy and immunotherapy in large, multi centric trial settings.

## Introduction

Esophageal cancer is the eighth most common cancer in the world, with an estimated 604,100 new cases and the sixth leading cause of cancer-related mortality in the world with 544,076 deaths reported as per GLOBOCON 2020^1^. However, there is a large geographic variation in its epidemiology, with the age-standardized incidence rate of esophageal cancer being 18.2/lac and 9.4/lac in Eastern Asia and Southern Africa respectively and only 1.5/lac in Central America and Western Africa^2^. Though the incidence rate in South-East Asian countries including India is 3.74/lac and 0.72/lac among male and female population respectively, it is showing an increasing trend^2^. With an incidence of 63,180 cases in 2020, it is the most common gastro-intestinal cancer in India^3^. The two most common types of esophageal cancer are squamous cell carcinoma (SCC) and adenocarcinoma (AC), which has different etiologies, biological characteristics and geographic distributions^4^. The etiological factors for esophageal squamous cell carcinoma include use of tobacco, alcohol, poor diet deficient in vitamins, indoor pollution, and consumption of hot beverages; whereas esophageal adenocarcinoma has been found to be associated with gastro-oesophageal reflux disease, obesity, tobacco use, and genetic risk factors^5-7^.

Overall, esophageal carcinoma is associated with poor survival, and with a mortality rate (5.6 per 100 000) that is close to the incidence rate (6.3 per 100 000)^2^. The incidence and mortality of esophageal cancer are further higher in developing countries compared to developed countries because of poor lifestyle, lack of adequate infrastructure for the implementation of cancer screening, and limited access to standard cancer care for the general population^8, 9^. The low 5-year survival rate of 15%–25% for oesophageal cancer is mainly due to the advanced stage at diagnosis, as majority of the patients present with unresectable or metastatic disease^10, 11^. Though surgery is the treatment of choice for esophageal cancers, radiotherapy alone has been used as an alternative local treatment for esophageal cancer not amenable to surgery, but the outcome is unsatisfactory due to poor local control and distant metastasis^12,13^. The addition of chemotherapy to radiotherapy is synergistic as it not only enhances the local effects of radiation, but it also eliminates micro metastases and thus decreases distant metastasis^14^. Based on the landmark Radiation Therapy Oncology Group (RTOG) trial 85-01, which showed that the use of Cisplatin, 5-FU and concurrent radiation in esophageal cancer resulted in a 5-year survival rate of 26%, concurrent chemo radiotherapy has been used as the standard treatment for unresectable locally advanced esophageal cancer^15^. To further improve up on the survival rates, different chemotherapy and targeted therapy agents have been tried in combination with radiotherapy in various phase II and III trials ^16^.

As a result, there are a number of treatment options available for the non-surgical treatment of esophageal cancer. Patients especially those from low socio-economic strata present with long standing dysphagia resulting in weight loss and poor general condition, which preclude them from receiving the standard protocol of chemo radiotherapy. There is a general perception among oncologist that the clinical outcomes of patients with esophageal cancer in routine clinical care differ from those in randomized controlled trials. Besides, there are limited data regarding real-world clinical practice in the field of esophageal cancer from our part of the world. So, we conducted this study with an aim to provide real world data on treatment and outcome in our patients with esophageal cancer.

## Materials and Methods

### Study population

A total of 128 patients of esophageal cancer who were treated in the department of Radiotherapy, Mahavir Cancer Sansthan, Patna from 1^st^ January 2016 to 31^st^ December 2017 were evaluated for this retrospective study. Given the retrospective nature of the study, approval from the Institutional Ethics Committee was not required as a part of our institutional protocol, and the need for obtaining written informed consent was also waived. The inclusion criteria for the study included patients with histologically confirmed squamous cell carcinoma (SCC) or adenocarcinoma (AC) of the esophagus which were inoperable or unresectable and received radiotherapy or concurrent chemo radiotherapy with a radical intent. Patients were defined as unresectable when they had extensive disease (T4, extensive and bulky Nodes) or technical unresectable tumor (high cervical localization). Patients were defined as inoperable when co-morbidities excluded them from surgery. Patients were excluded from the study if they had received palliative radiotherapy because of metastatic disease or poor performance status of the patient or with other histology (non SCC or AC). [Flowchart]

### Data Collection

Data regarding patients’ age, histology, location, pre-treatment imaging, disease stage, treatment details, compliance and response to treatment and status at last follow-up were retrieved from their file. The histological subtypes (SCC and AC) and grades were assigned based on the WHO classification. Anatomical location was defined as upper-third including cervical part (15–25 cm from the incisors), middle-third (>25–30 cm), and lower-third cancer (>30 cm) based on the epicenter of the esophageal tumor. If multiple lesions were present, the location was defined based on the location of the bulk of the lesion. The cancer stage at the initial diagnosis was defined according to the 7th edition of American Joint Committee on Cancer (AJCC) TNM staging system. The staging was based on findings from barium swallow, endoscopy, chest x-ray, ultrasonography of abdomen, computed tomography, and positron emission tomography, as available. Radiotherapy was delivered on linear accelerators by three-dimensional conformal radiotherapy (3DCRT) technique or intensity modulated radiotherapy (IMRT) technique. The target volume and organs at risk were contoured on contrast-enhanced CT images fused with positron emission tomography images whenever available. The gross tumor volume (GTV) was contoured as the contrast-enhancing tumor along with endoscopy correlation. The clinical target volume (CTV) was contoured as GTV expanded by 3 to 4 cm superiorly and inferiorly along the length of the esophagus and cardia and 1-cm radial expansion. The nodal CTV was contoured as 0.5 to 1.5 cm expansion from the nodal GTV. The celiac axis was covered for tumors of the distal esophagus or gastroesophageal junction. Supraclavicular nodes were included for the upper thoracic esophageal tumors. CTV was expanded by 1.0 cm to generate planning treatment volume (PTV). The boost CTV was created by 1 to 1.5 cm expansion from the initial GTV and further expanded by 1cm to create the boost PTV. Initial PTV was treated to a dose of 41.4 to 45 Gy at 1.8 Gy per fraction, followed by a cone-down boost to a total dose of 50.4 to 63 Gy in 1.8 Gy per fraction. Concurrent chemotherapy consisted of either single agent Cisplatin 40mg/m^2^ or a combination of paclitaxel 50 mg/m2 and carboplatin area under the curve (AUC) 2 administered intravenously (IV) once weekly for five to seven cycles along with RT.

### Follow-up

After treatment completion, patients were seen at 6 to 8 weeks for the first follow-up, then at 3-month intervals for the first 12 months, then every 6 months up to year 3, and annually thereafter. A detailed clinical evaluation and physical examination was done on every follow-up. CT scans of the chest and upper abdomen were advised every 6 months for 3 years and annually thereafter. Other studies, such as endoscopy and PET scanning, were done only if clinically indicated.

### Treatment Response

Treatment response was assessed by detailed clinical and physical examination and using CT scan and endoscopy at 6 to 8 weeks after completion of treatment. PET-CT was used if clinically indicated and feasible for the patient. Patients with no clinical evidence of disease and normal CECT scan or PET-CT scan at their last follow-up or 3 years after their diagnosis were classified as loco regionally controlled (LRC). Patients who had persistent or worsening of symptoms and showed disease on imaging and endoscopy at first follow-up were classified as having residual disease (RES). Patients having an initial complete response to treatment but developing recurrence as defined by clinical symptoms and signs, suspicious endoscopic findings, combined with imaging findings on CECT scan or PET/CT scan, located at the site of the primary tumor and/or at the site or regional lymph nodes was classified as loco regional failure (LRF). Histological confirmation was done whenever feasible and in all cases with suspicious finding on CECT, PET-CT or endoscopic examination. Patients showing metastases to non-regional lymph nodes, distant organs with or without loco regional disease were classified as distant metastases (DM). Site of distant metastases was documented to study the pattern of distant failure.

### Statistical Analysis

All the relevant data was entered in Microsoft excel sheet. The disease status of the patients was entered until death, local recurrence, or their last follow up. Continuous and categorical variables were summarized by descriptive statistics using online calculator. Continuous data was analyzed in terms of range, mean with standard deviation and median with inter quartile range. Categorical data was expressed in percentage for comparison.

## RESULTS

A total of 100 esophageal cancer patients were included in the study. The mean age of the patient was 60.24 ± 11.45 year with a range from 29 to 85 years. Majority of the patients (60%) were male with a male to female ratio of 1.5:1. Upper one-third of esophagus was the most common site affected followed by lower and middle third consisting of 40%, 36% and 24% of the cases respectively. Squamous cell carcinoma was the predominant histology, reported in 83% of cases with adenocarcinoma in remaining 17%. Most of the patient had grade II carcinoma, which was seen in 54% of the cases followed by 26% grade I, 11% grade III and 9% with unknown grade. CECT scan was done in 59% of the patients, while PET-CT scan was reported in only 17% of the patients. Remaining patients underwent barium swallow, chest x-ray and ultrasonography of abdomen before starting the treatment for esophageal cancer. T and N staging could be done in only 76% of these patients with 69% showing T3-4 stage and 7% with T1-2 stage. Further, 32% of the patients were node negative on imaging and 44% showed nodal disease on CECT or PET-CT scans. [Table1]

**Table 1:** Showing baseline characteristics of the patient cohort

| Parameter | Number (%) |
| --- | --- |
| **Age** |  |
| Range | 29 – 85 years |
| Mean ± S.D | 60.24 ± 11.45 |
| Median (IQR) | 62 (18) |
| **Sex** |  |
| Male | 60 (60%) |
| Female | 40 (40%) |
| Ratio | 1:5 |
| **Site** |  |
| Upper | 40 (40%) |
| Middle | 24 (24%) |
| Lower | 36 (36%) |
| **Histology** |  |
| Squamous cell carcinoma | 83 (83%) |
| Adenocarcinoma | 17 (17%) |
| **Grade** |  |
| **1** | 26 (26%) |
| **2** | 54 (54%) |
| **3** | 11 (11%) |
| **Unknown** | 9 (9%) |
| **Imaging** |  |
| **CECT** | 59 (59%) |
| **PET-CT** | 17 (17%) |
| **Others** | 24 (24%) |
| **T status** |  |
| **T1-2** | 7 (7%) |
| **T3-4** | 69 (69%) |
| **Unknown** | 24 (24%) |
| **N status** |  |
| **N0** | 32 (32%) |
| **N+** | 44 (44%) |
| **Unknown** | 24 (24%) |

CECT = Contrast enhanced computed tomography, PET-CT = Positron emission tomography

Regarding treatment details, 15% of the patients received Neoadjuvant chemotherapy before radiotherapy. The most common regimen used was 3 weekly Paclitaxel (175mg/m2) and Carboplatin (AUC 5) in 11 patients. 5-FU with Cisplatin was used in three patients while one received 5-FU, Docetaxel and Cisplatin. Fifty four percent of the patients received concurrent chemotherapy with radiation. The most common drug used was weekly Cisplatin (40mg/m2) followed by weekly Paclitaxel (50mg/m2) and Carboplatin (AUC 2). Single agent concurrent Cisplatin was used in 33% of patients and 21% received doublet chemotherapy with Paclitaxel and Carboplatin. Majority of the patients, 76% received radiation by 3DCRT technique and remaining 24% by IMRT technique. The radiation dose varied from 45Gy to 63Gy, with a median dose of 59.4Gy. Seventy five percent of the patients received a dose higher than 50.4Gy. Ninety percent of the patients completed their planned radiotherapy protocol, while 10% defaulted during radiation because of toxicities, worsening of symptoms or personal reasons. [Table 2]

**Table 2:** Showing treatment pattern and compliance of the patient cohort

| **Parameter** | **Number (%)**<br>**(N = 100)** |
| --- | --- |
| **Neoadjuvant Chemotherapy** | 15 (15%) |
| **Paclitaxel + Carboplatin** | 11 (11%) |
| **Others** | 04 (4%) |
| **Concurrent Chemotherapy** | 54 (54%) |
| **Cisplatin** | 33 (33%) |
| **Paclitaxel + Carboplatin** | 21 (21%) |
| **Radiation Technique** |  |
| **3DCRT** | 76 (76%) |
| **IMRT** | 24 (24%) |
| **Radiation Dose** |  |
| **Range** | 45 – 63Gy |
| **Mean ± S.D** | 57.84 ± 5.72 |
| **Median (IQR)** | 59.4Gy (10.8) |
| **50.4Gy** | 25 (25%) |
| **> 50.4Gy** | 75 (75%) |
| **Treatment Compliance** |  |
| **Completed** | 90 (90%) |
| **Defaulted** | 10 (10%) |

3DCRT = 3 dimensional conformal radiotherapy technique, IMRT = Intensity modulated radiotherapy technique

Among 90 patients who completed their planned radiotherapy, 5 patients had no follow-up. After a mean follow up of 10.98 months (range 3 -58 months, median FU 7 months), a total of 72 patients (80%) were alive with 29 (32.22%) of them having no evidence of disease. Loco-regional failure was seen in 19 (21.11%) patients, while 5 (5.55%) of them had a residual disease. Further, 19 (21.11%) of the patients were alive but with distant metastases. Out of 13 (14.44%) patients who had died, 10 (11.11%) had distant metastases, 2 (2.22%) had complications and 1(1.11%) had a loco regional failure with tracheo-esophageal fistula. [Table.3]

**Table 3:**
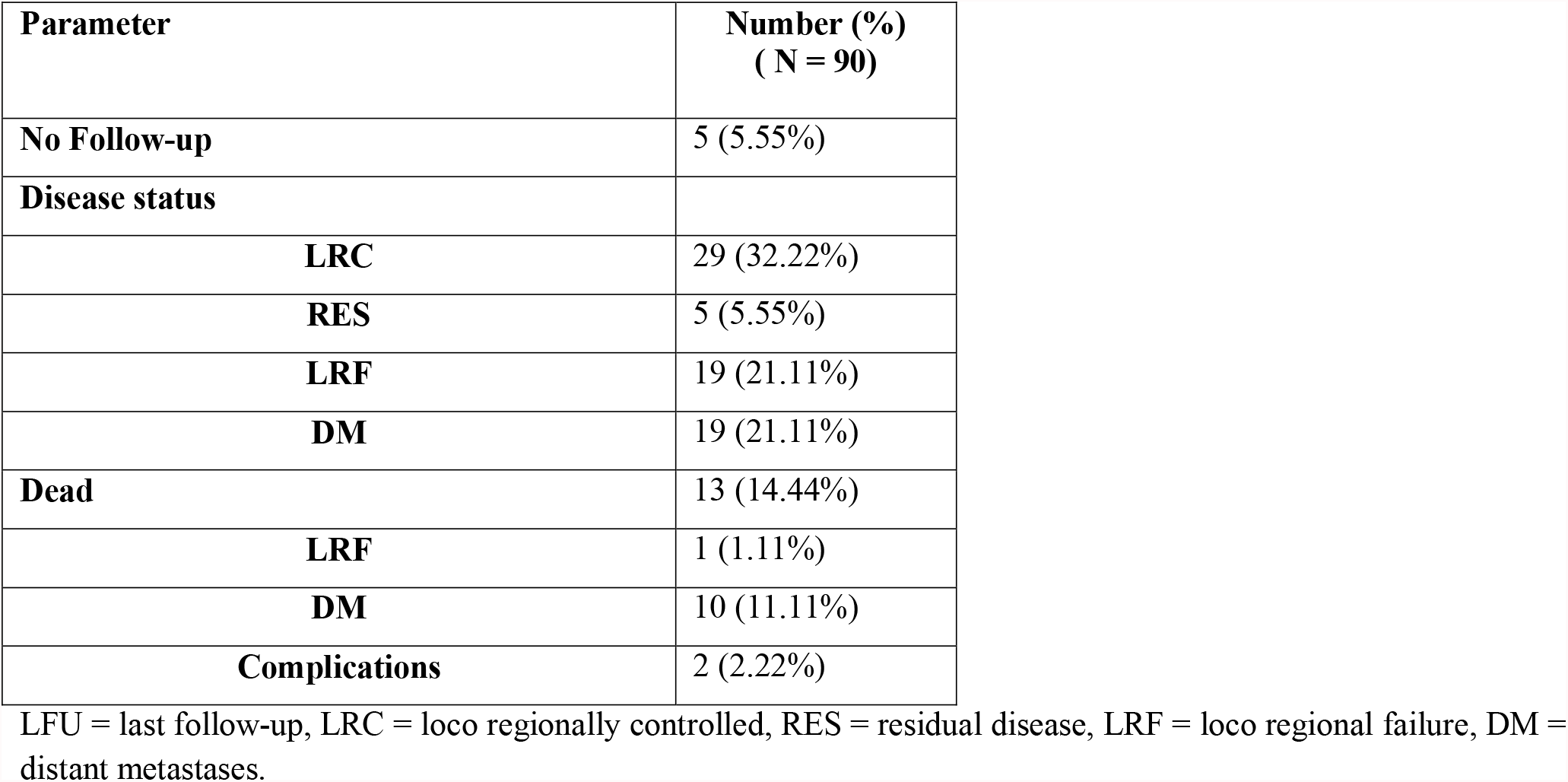
Showing treatment outcome of the patient cohort

| **Parameter** | **Number (%)**<br>**( N = 90)** |
| --- | --- |
| **No Follow-up** | 5 (5.55%) |
| **Disease status** |  |
| **LRC** | 29 (32.22%) |
| **RES** | 5 (5.55%) |
| **LRF** | 19 (21.11%) |
| **DM** | 19 (21.11%) |
| **Dead** | 13 (14.44%) |
| **LRF** | 1 (1.11%) |
| **DM** | 10 (11.11%) |
| **Complications** | 2 (2.22%) |

LFU = last follow-up, LRC = loco regionally controlled, RES = residual disease, LRF = loco regional failure, DM = distant metastases.

A subset analysis of patients with distant metastases showed liver to be the most common site of metastases followed by lung, bone, brain, and non-regional lymph nodes in 10(11.11%), 8(8.88%), 7 (7.77%), 2(2.22%), 2(2.22%) patients respectively. One patient had bilateral ovarian metastases and another showed multiple metastatic subcutaneous nodules over trunk and upper back. Three patients with lung metastases also had malignant pleural effusion. One patient had both liver and lung metastases, whereas the patient with ovarian metastases also had bone metastases. [Table. 4]

**Table 4:** Showing sites of distant metastases of the patient cohort

| **Site** | **Number (%)**<br>**(N = 90)** |
| --- | --- |
| **Liver** | 10 (11.11%) |
| **Lung** | 8 (8.88%) |
| **Bone** | 7 (7.77%) |
| **Brain** | 2 (2.22%) |
| **Supra clavicular lymph node** | 2 (2.22%) |
| **Ovary** | 1 (1.11%) |
| **Sub cutaneous nodules** | 1 (1.11%) |

## Discussion

In this study, we have retrospectively analyzed the clinical characteristics, treatment and outcome in patients with unresectable or inoperable esophageal cancer. The mean age of our patient cohort was 60.24 years (range 29-85 years) with a male to female ratio of 1.5:1. Population based data reveal esophageal cancer to be a disease of the elderly with the peak of incidence in the sixth decade of life^17^. The mean age of patients suffering from esophageal cancer in Asian countries has been reported to be in range of 51–60 years^18^. Further, esophageal carcinoma has a predilection towards males, affecting males 2-4 times more frequently as compared to females worldwide^19^. Chokshi et al in their epidemiological analysis of esophageal cancer in India reported a mean age of 54.83 years (range 25–89 years) with a male to female ratio of 1.67:1^20^. Another study from north-west India showed a mean age of 54.1 years and a male to female ratio of 1.15:1^18^. The reason for higher prevalence in females in India than that reported in studies from Western population needs to be identified. A study from Punjab found poor nourishment and consumption of hot beverages to be linked to SCC carcinogenesis among female patients^21^.

Our study showed upper esophagus as the most common site affected followed closely by lower and middle esophagus consisting of 40%, 36% and 24% of the cases respectively. This finding is in contrast with that of other study from western India where the most common location was mid esophagus^20^. However, a recent study from eastern India reported upper esophagus to be the most common site seen in 47.2% of the patients^22^. Squamous cell carcinoma was the predominant histology reported in 83% of our cases with 54% of them having grade II carcinoma. Various Indian studies have reported Squamous cell carcinoma to be present in about 80% of all cases of esophageal cancer, similar to that seen in our patient cohort^23^. Though Squamous cell carcinoma has also been reported to be the predominant histology worldwide, there has been a rapid rise of adenocarcinoma in western countries in recent years^24^.

A contrast enhanced CT scan (CECT) was the most common imaging modality used for staging in our patients. About 60% of our patients underwent CECT scan and only 17% underwent PET-CT scan for staging. Based on these imaging modalities, 69% of our patients had T3-T4 disease and only 7% had T1-T2 disease. Regarding nodal status, 44% of our patients had radiologically involved nodes and 32% had no significant lymhadenopathy on imaging. Remaining 24% of the patients were assigned an unknown stage as they had either received neo-adjuvant chemotherapy based on results of endoscopy, biopsy and, barium swallow at other centre before being referred for radiotherapy or were not affording/willing for further investigations and wanted treatment. Endoscopic ultrasound (EUS) helps to delineate the various layers of the esophagus and is considered superior to CECT scan in regard to loco regional staging of tumor invasion and lymph nodal involvement^25^. However, EUS has its own limitations and may not be technically feasible in obstructive growths^26^. In the Indian setting, because of the advanced nature of disease at presentation as well as its limited availability in many centers, the routine use of EUS is debatable^23^. Therefore, a CECT scan of the thorax and upper abdomen is widely accepted as the preferred modality of staging for cancer of the esophagus in the Indian setting. Positron emission tomography (PET) provides additional staging information, especially when combined with a CT (PET-CT) and is the best modality for detecting distant metastasis^27^. A recent study from Tata Memorial Hospital, Mumbai, evaluating the role of PET-CT in esophageal cancer patients reported detection of unsuspected metastatic disease in 16% patients^28^. However, the cost, availability, and the high false-positive rate due to infections such as tuberculosis are the practical difficulties in routine use of PET-CT in most cancer centers of our country^29^.

In our study, the treatment received included Neoadjuvant chemotherapy followed by concurrent chemo radiotherapy, concurrent chemo radiotherapy or radiotherapy alone. Among patients receiving chemotherapy 15 patients (15%) received it as neo adjuvant therapy. Ten (66.67%) of these patients also received concurrent chemo radiotherapy. Forty four (44%) of patients received upfront concurrent chemo radiotherapy, whereas 46% of patients received only radiotherapy as a definitive treatment.

Definitive concurrent chemo radiotherapy has been recommended as the standard non-surgical treatment for patients with oesophageal cancer. Based on the landmark RTOG 85-01 trial, Cisplatin and 5-FU along with radiation has remained the standard protocol for years. The above trial reported a 5-year survival rate of only 26% with a median survival of 14 months and grade 3–4 adverse reactions in 46% of the patients. Besides, this protocol uses continuous infusion of 5-FU for 4 days which requires admission causing logistic issues for the patients^15^. To further improve on the results and decrease the toxicities several chemotherapy combinations have been used concurrently with radiation therapy. These include trials combining radiation with paclitaxel and cisplatin, 5-FU and oxaliplatin, irinotecan and cisplatin, docetaxel cisplatin and 5-fluorouracil (DCF), Cisplatin and capecitabine, cetuximab with cisplatin and capecitabine^30-35^. The CROSS trial used a novel regimen of weekly paclitaxel and carboplatin concurrently with RT as a neo-adjuvant therapy followed by surgery and reported an unprecedented median overall survival of over 4 years^36^. Since then, several oncologists have successfully explored the use of this regimen in the definitive chemo radiation. A study by Noronha et al in Indian patients showed that concurrent chemo radiotherapy with weekly paclitaxel and carboplatin is well tolerated in Indian patients. The study showed a 3 year survival rate of 39% with an objective response rate of 48.6%, median progression free survival of 11 months and manageable toxicities^37^. In our study, concurrent paclitaxel and carboplatin was used in only 21% of the patients and another 33% received concurrent single agent cisplatin along with radiation. Use of weekly Cisplatin as radiosensitizer is a well established drug incorporating concurrent chemotherapy in radical treatment of squamous cell carcinoma of cervix and head and neck cancers^38,39^. Because of the ease of administration and better treatment compliance, weekly Cisplatin has also been used in definitive chemo radiation for esophageal cancer patients^40^. Ahmed et al reported a median OS of 15.2 months and 2 year OS of 42.6% in esophageal cancer patients treated with concurrent weekly cisplatin and radiation which was similar to that reported in the FFCD 9102 and Cisplatin-5 FU arm of Prodige5/Accord17^31,41-42^. The study using single agent cisplatin also had a lower incidence of grade 3 or higher toxicities and all were hematological. Li et al in their multicenter retrospective analysis comparing the therapeutic effects of single-agent and double-agent concurrent chemo radiotherapy in patients with unresectable esophageal squamous cell carcinoma suggested that single therapy is not inferior to dual therapy especially in the elderly patients^43^.

Another protocol used to improve on the survival of esophageal cancer patients treated with definitive chemo radiation is the use of induction or neo adjuvant chemotherapy before starting chemo radiation. The underlying rationale is to reduce the bulk of primary tumor and control distant micro metastases. Induction with DCF followed by concurrent CRT using carboplatin has been reported to have a CR rate of 16 % and median overall survival of 10.8 months in a trial by Chiarion-Sileni et al^44^. The phase II COSMOS trial conducted by Yokota et al combining induction chemotherapy using docetaxel plus cisplatin and 5FU (DCF) followed by radical CRT, and conversion surgery, when feasible, reported promising results with a 3-year overall survival of 46.6% at a median follow-up of 39.3 months in locally advanced unresectable esophageal cancer^45^. Based on this result, the JCOG has started a phase III trial (JCOG1510) investigating the efficacy of induction chemotherapy using DCF followed by conversion surgery and/or radical CRT in patients with locally advanced unresectable squamous-cell carcinoma of thoracic esophagus. The primary endpoint of this ongoing trial is overall survival with progression-free survival, complete response rate of concurrent chemo radiation, and toxicities as the secondary end-points^46^. Though efficient, DCF protocol has been associated with considerable toxicities. Further, neoadjuvant treatment with carboplatin and paclitaxel-based chemotherapy produced a 27.9% pathologic complete response rate in patients with resectable esophageal cancer, according to results of the NEOSCOPE trial^47^. Most of the patients in our study received Paclitaxel and Caboplatin in the neo adjuvant setting keeping in view good response to doublet chemotherapy and a higher toxicity and cost associated with the triplet regimen.

However, till date the long term results of definitive chemo radiation with or without induction chemotherapy show poor survival and multiple new treatment strategies are being tried. As a result, the standard practice of concurrent chemo radiation in carcinoma esophagus varies substantially throughout the world and even in our country. Besides, a considerable number of patients undergo a single modality of treatment as seen in our study because of the fear that multimodality treatment may not be tolerated by the generally frail patients with esophageal cancer and also because of their advanced age at diagnosis with inadequate nutritional support. Similar to our study, a meta-analysis by Zhu et al of concurrent chemo radiotherapy for advanced esophageal cancer showed that 46% of the patients received only radiotherapy. The overall response rate was 93.4% for concurrent chemo radiotherapy and 83.7% for radiotherapy alone (P= 0.05). However, CCRT arm showed a better 3-year and 5-year survival with an increased incidence of acute toxicities^48^.

Another important issue in the non-surgical treatment of esophageal cancer is the dose of radiotherapy. On the basis of results from RTOG 94–05, 50.4Gy has been accepted as standard dose in western countries and also recommended by NCCN guideline in both neo-adjuvant and radical setting^49,50^. Based on the theory of radiation biology, 50.4Gy is just adequate to control microscopic cancer cell, but inadequate to control a gross tumor lesion. A radiation dose more than 60Gy or even nearly 100Gy is required to control and cure a gross solid tumor^51^. Further, with the clinical application of more precise radiation techniques such as IMRT, interpretation about the results of RTOG 94–05 should be different. A pooled analysis from Song et al. showed that a higher radiation dose could improve clinical outcomes without significantly increasing radiation-related toxicities^52^. On this basis, a radiation dose of 60 to 66Gy is used in many Asian countries including Japan^53^. At the time of patient accrual for the current study, there were several ongoing clinical trials evaluating dose-escalation strategies using state-of-the-art RT technology to improve loco regional control, including the SCOPE2 trial (50Gy versus 60Gy), CONCORDE trial (50Gy versus 66Gy), and Art-Deco trial (50.4Gy versus 61.6Gy)^54-56^. In our study, 25% of the patients received a radiation dose of 50.4Gy and remaining 75% received radiation dose more than 50.4Gy with a median dose of 59.4Gy. A radiation dose of more than 50.4Gy was well tolerated as 90% of our patients completed the planned radiation protocol. Another reason for the use of high median dose of radiation in our study was the large number of patients with upper esophageal cancer. It is believed that the biological behavior of upper esophageal cancer differs from those at the mid and lower esophagus, because they are mostly squamous-cell histology with local invasiveness and less prone to distant metastasis, and that they should be treated like head and neck cancer. Wang et al analyzed the treatment and outcome of patients with cervical and upper thoracic esophageal cancer and found that radiation dose was the only independent factor associated with improved local control and overall survival. They concluded that OS and DFS were significantly higher in patients who had received a radiation dose of greater than or equal to 50Gy than in those who had received a dose of less than 50Gy^57^.

The survival rate of 32.22% at the end of 3 years seen in our study is similar to that reported in literature. Various studies have shown a 3 year survival rate of 20% to 45% in esophageal cancer treated with concurrent chemo radiation^16^. An Indian study on clinical outcome in definitive concurrent chemo radiation with weekly paclitaxel and carboplatin for locally advanced esophageal cancer reported 1-year, 2-year, and 3-year survivals of 70%, 47%, and 39%, respectively^37^. However, our study showed distant metastases as the most common site of failure followed closely by loco regional recurrence. This is in contrast to most of the studies which have shown loco regional failure as the most common site of treatment failure^58,59^. This could be because of the fact that baseline PET-CT and CECT scan was available in only 17% and 59% of patients respectively. So, there are chances that asymptomatic distant metastases present at the time of diagnosis were missed. Another reason could be the use of chemotherapy either in neo-adjuvant or concurrent form in only 56% of our patients. Liver was the most common site of distant metastases seen in our study followed by lung, bone and brain. The pattern reported is similar to that seen in the study by Wu et al analyzing pattern of distant metastases in patients with de novo stage IV esophageal cancer at diagnosis identified using the Surveillance, Epidemiology, and End Results database^60^.

Before we conclude, it is important to describe the limitations of this study. Being a retrospective analysis, only documented details were available for evaluation. Being a single center study, the sample size was small and heterogeneous. Because of non-availability of PET-CT scan and even CECT scan in few patients, the staging was inadequate in few patients. The patients were treated with a varied combination of chemotherapy and radiation dose. This precluded any subset analysis for predictors of loco regional and distant control in esophageal cancer patients treated with radiation with or without chemotherapy. Nonetheless, this real world data will surely bring forward the issues and outcomes of esophageal cancer patients treated outside clinical trials and may help in designing new studies.

## Conclusion

The study showed that squamous cell carcinoma remains the predominant histology in our population with upper esophagus as the most common location. Esophageal cancer continues to be a disease of the elderly. Inadequate nutritional support and presence of co morbidities remains a hindrance for a uniform treatment protocol using concurrent chemo radiation. Both distant metastases and loco regional failure continues to be a matter of concern. Routine use of new imaging modalities like PET-CT scan must be done for adequate staging of these patients to rule out distant metastases at the time of diagnosis. Further improvement in local control must be evaluated by either radiation dose escalation or novel combinations with chemotherapy and immunotherapy in large, multi centric trial settings.

## Data Availability

All data produced in the present work are contained in the manuscript

## FLOWCHART

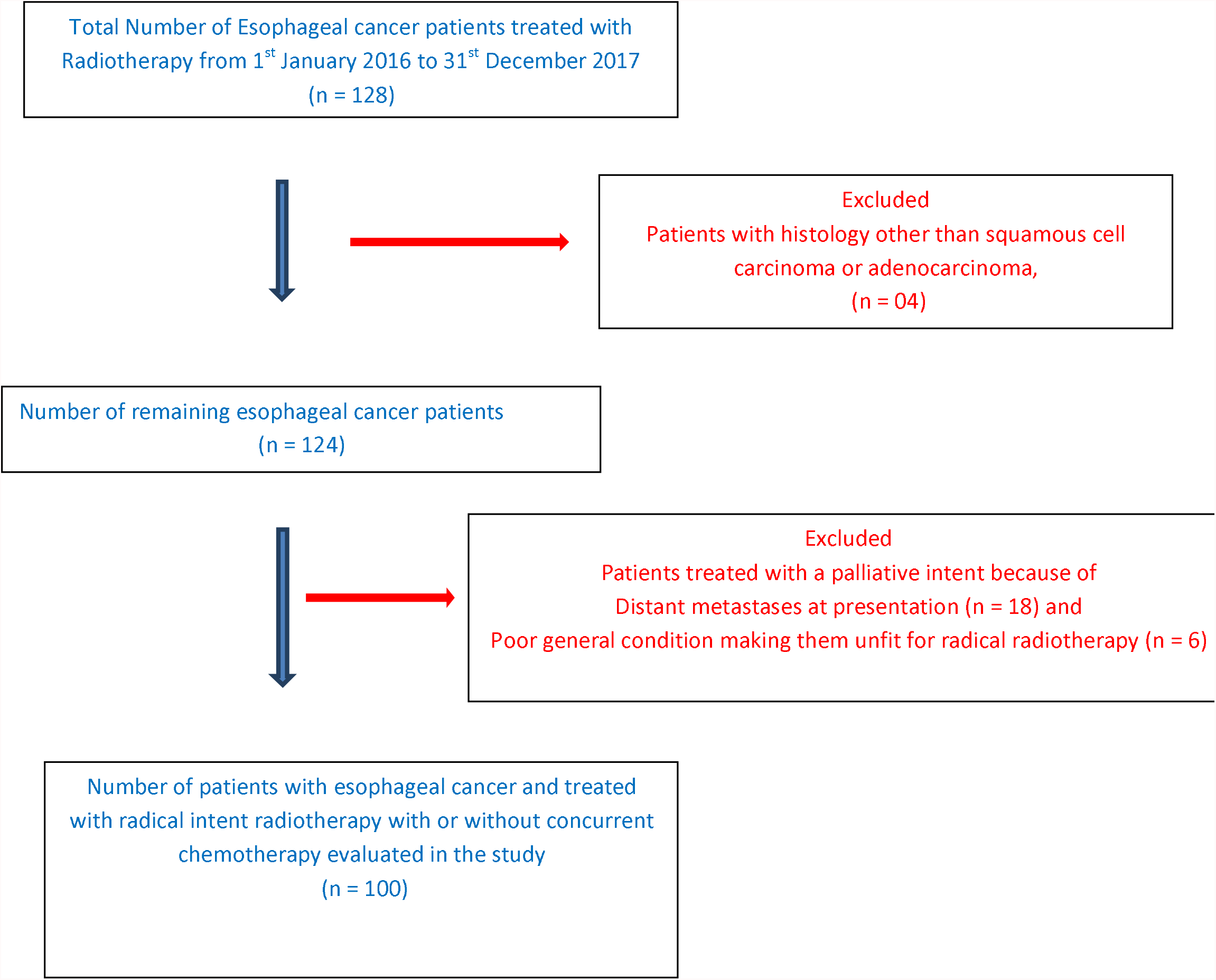

